# Changing Physician Performance: A Systematic Review and Meta-Analysis (75 years 1950-2024) of the Effect of Continuing Medical Education Strategies, Continuous Professional Development and knowledge Translation

**DOI:** 10.1101/2025.02.06.25321832

**Authors:** Ebrahim Abbasi

**Affiliations:** Medical Education Department, Medical Education Development Center, Shiraz University of Medical Sciences, Shiraz, Iran; Department of Medical Entomology and Vector Control, School of Health, Shiraz University of Medical Sciences, Shiraz, Iran

**Keywords:** Physician Performance, Continuing Medical Education Strategies, Continuous Professional Development, knowledge Translation, Systematic Review

## Abstract

**Introduction:** Medical education has always been considered an essential component for improving the knowledg and performance of physicians and medical assistants. However, this education must be carried out continuously t update the knowledge of physicians and improve their performance. Various strategies are used to achieve this goal, the most important of which are CME, CPD, and CBME strategies. This study aimed to investigate the effect of using these strategies on improving the performance of physicians in the world using a systematic review method.

**Methods:** This study was conducted using a systematic review method on the effect of educational strategies o improving the performance of physicians in the world. Accordingly, all relevant articles published between 195 and 2024 were extracted and reviewed through a search in the scientific databases PubMed/Medline, Web of Science, ProQuest, and Scopus. The quality of the articles was assessed using the CONSORT checklist an Newcastle-Ottawa Scale.

**Results:** twenty articles conducted between 1993 and 2024 were included in the systematic review process. According to the findings, providing continuing education based on CME, CPD, CBME, and knowledge translatio strategies positively affected the knowledge, skills, and performance of physicians and medical assistants i providing health services. This can be used to train medical students and practicing physicians on an ongoing basis.

**Conclusion:** Medical education using CME, CPD, and CBME strategies can improve physicians’ knowledge and performance, and subsequently increase job satisfaction, self-efficacy, communication skills, medical knowledge, performance, and skill. However, individual, structural, and content limitations that can prevent success in learnin and training programs should be considered and addressed.

## INTRODUCTION

Medical education focuses on providing training and practical experience to physicians, medical staff, and students. The goal of this type of education is to enhance knowledge, learning, performance, and interaction with patients and other members of the community. Medical education can be conducted in various settings, including hospitals, clinics, universities, and educational centers (1, 2). To improve the quality of health services and care provided to patients by providing clinical and practical education (3). Today, various approaches are used to educate medical groups, including Continuing Medical Education (CME), Continuous Professional Development (CPD), and Community-Based Medical Education (CBME) (4, 5).

Continuing medical education (CME), as an essential component of medical education systems, provides lifelong learning opportunities to individuals studying or working in the medical field and improves and ensures the quality of their performance (6). Continuing education (CE) maintains competence and develops knowledge and science in new areas of medicine for healthcare professionals. CME system activities can be carried out in the form of online programs, live and web-based educational programs, written publications, simulations, conferences, scientific meetings, and practical practice-based skills. These programs or contents can be presented or developed by experienced professors in specialized fields (7, 8). Continuing professional development (CPD) is an effective strategy for maintaining and developing knowledge and skills in the field of medical professions. This system can strengthen skills, develop them to a new level, and expand their job roles (9). Many companies and organizations constantly prioritize CPD due to the need to develop and improve the job performance of their workforce. CPD activities can include workshops, conferences, seminars, and training courses, as well as informal approaches such as reading, work-based learning, or variable mentoring (10, 11). Community-based medical education (CBME) is another approach used to educate, empower, and improve the knowledge and practice of medical staff, emphasizing the importance of community participation in the educational process (2). The goal of CBME is to empower individuals and communities to actively shape educational experiences and outcomes (12). This approach promotes the participation of communities, families, and medical staff in health care, and improves their knowledge, practice, and well-being (13). Community-based medical education for physicians and medical residents, by focusing on curriculum design, clinical education, and clinical practice, can promote the social accountability of the health sector and also create the ability to adapt to different cultures (14).

Various countries in the world, including European and American countries, emphasize the use of strategies to improve the knowledge and performance of physicians, including CME, CPD, and CBME, and in many of them, participation in educational programs based on these approaches has been mandatory for physicians (6, 7). In general, today, attention has been paid to the use of the aforementioned strategies in the form of conferences, congresses, and web-based programs to improve the performance of physicians and medical assistants (4). However, some studies have mentioned contradictions regarding the use of strategies and their associated limitations. Accordingly, this study was conducted to investigate the use of CME, CPD, and CBME strategies to improve the performance of physicians and medical assistants in the world using a systematic review method.

## MATERIAL AND METHODS

This study was conducted as a systematic review of physician practice change strategies worldwide according to the Preferred Reporting Items for Systematic Reviews and Meta-Analysis (PRISMA) guidelines (15).

This research has been registered in the International Prospective Register of Systematic Review (PROSPERO) with the CRD42025649098.

### Search strategy

Searching for articles in the international databases PubMed/Medline, Web of Science, ProQuest, and Scopus without time limits until the end of 2024 using the keywords Doctor’s Assistant, Physicians’ Extender, Physicians’, Physician Assistant, Continuing Medical Education, Education, Continuing Medical, Medical Education, professional development, staff development, professional training, workplace training, professional education, ongoing training, continuing professional development, Biomedical Translational Science, Translational Sciences. Translational Medicine, Knowledge Translation was performed individually and in combination using the OR and AND operators in the title, abstract, and full text, and all relevant articles were extracted.

### Inclusion and Exclusion Criteria

Articles with the following criteria: 1- Articles on changing the performance of physicians and medical staff; 2- CME, CPD, and knowledge translation strategies were examined; 3- They were conducted on physicians and medical staff; 4- The outcome of the intervention was specified; and 5- They were of desirable quality were included in the study. Articles that did not meet the inclusion criteria were excluded from the study.

### Quality Assessment

Article quality assessment was performed for interventional studies using the CONSORT checklist (16) and for observational studies using the Newcastle-Ottawa Scale (NOS) checklist (17) according to the guidelines. To classify the quality of articles, a score of less than 50% was considered poor quality, a score of 50 to 75% was considered moderate quality, and a score of more than 75% was considered high quality. Articles with moderate to high quality were included in the study.

### Selection of Studies

A total of 35,581 articles were extracted in the initial search. Then, the articles were entered into Endnote software duplicates were identified, and 7175 articles were excluded due to duplication. In the next step, by carefully studying the titles and abstracts of the articles, 26167 articles were excluded due to their irrelevance to the study. Then, the full text of 2239 articles were reviewed, and 2219 articles were excluded from the study due to the lack of examination of educational interventions and their impact on physician performance. Finally, 20 articles were included in the systematic review process (Figure 1).

**Figure 1.**
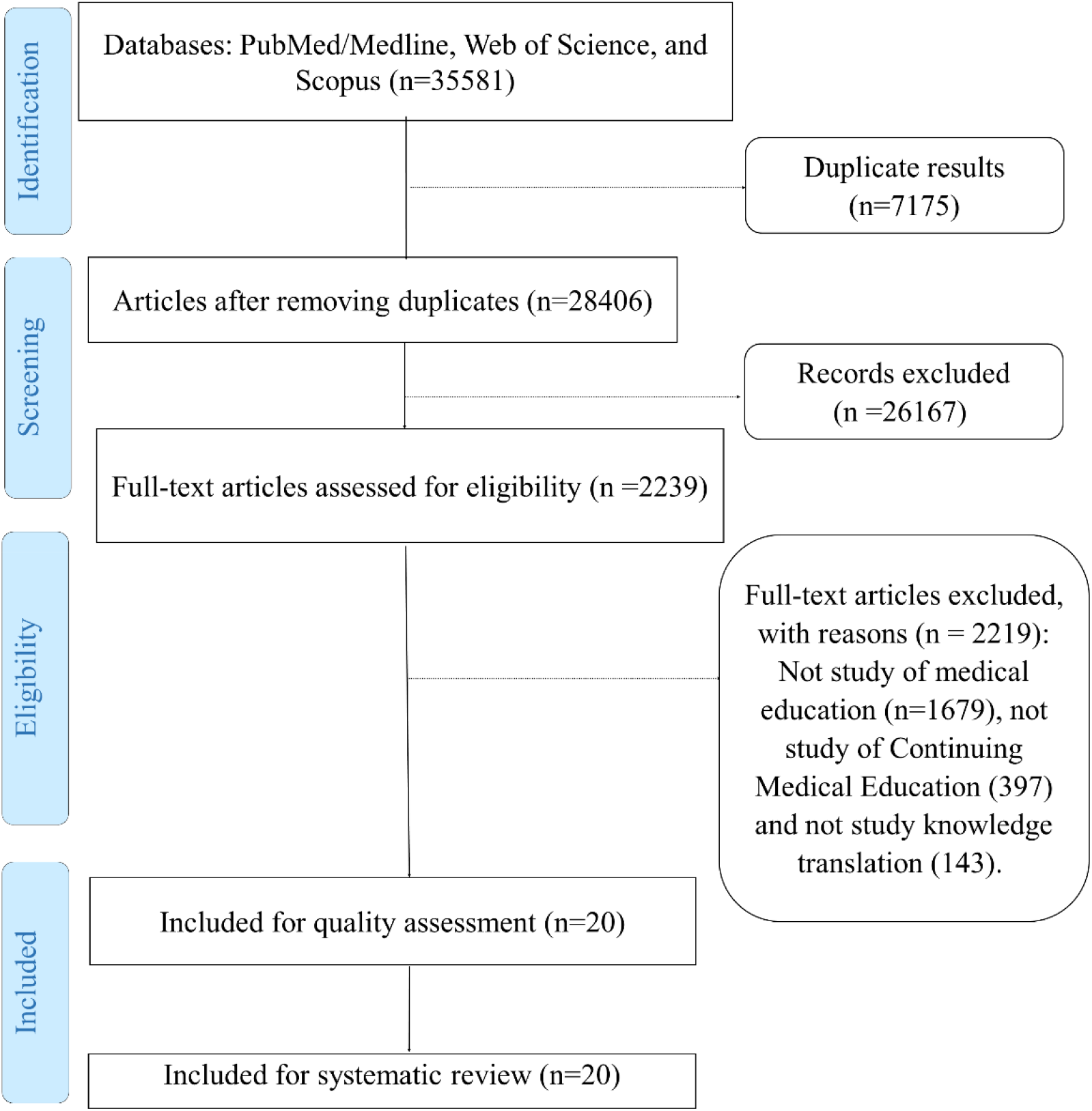
The PRISMA flow diagram of the search process

### Data extraction

Data extraction was performed independently by two researchers. Accordingly, the full text of the articles that met the inclusion criteria was first reviewed. If the articles were rejected by the two researchers, the reason was stated, and in case of disagreement between them, the article was reviewed by a third person. Data extraction was performed using a checklist that included the characteristics of the first author, the publication date of the article, sample size, strategy and type of intervention, and the outcome of the intervention.

## RESULTS

A total of 20 articles that were conducted between 1993 and 2024 were included in the systematic review process. 9 articles were conducted in the field of CME, 8 articles in the field of CPD, and one article each in the fields of CBME, RTME, and HPE. The characteristics of the articles included in the systematic review are presented in Table 1.

**Table 1.** Characteristics of articles included in the systematic review process.

| Author | Year of study | Place of study | Sample size | Quality assessment | Strategies |
| --- | --- | --- | --- | --- | --- |
| Wang H (18) | 2024 | China | 3954 | High | CME |
| Jayas A (19) | 2023 | America | 5926 | High | CME |
| Pott M (20) | 2021 | USA | 500 | Mild | CME |
| Gill C (21) | 2016 | Vietnam | 638 | High | CME |
| Shah N (22) | 2012 | Pakistan | 1200 | High | CME |
| Liljestrand P (23) | 2004 | Arizona, California, Hawaii, and Nevada | 731 | High | CME |
| Eickmann S (27) | 2024 | German | 13 | Mild | CME |
| Davis DA (25) | 1995 | World | 99 | High | CME |
| Levinson W (26) | 1993 | Portland | 62 | Mild | CME |
| Al Sheikhly D(28) | 2023 | Qatar | 281 | Mild | CPD |
| Tavares W (29) | 2022 | World | 4 | Mild | CPD |
| Kozikowski A (30) | 2021 | USA | 1388 | High | CPD |
| Whicker SA (31) | 2020 | USA | - | High | CPD |
| Breunig MJ (32) | 2020 | USA | 163 | High | CME and CPD |
| Alexander LM (33) | 2018 | USA | 75 | Mild | CPD |
| Howie N (34) | 2017 | England | 1000 | Mild | CPD |
| Rao R (35) | 2010 | England | 89 | Mild | CPD |
| Kulo V (36) | 2022 | USA and Canada | 20 | High | HPE |
| Miskeen E (5) | 2023 | Saudi Arabia | 72 | High | CBME |
| Jin R (37) | 2023 | China | 240 | High | RTME |

The use of educational methods to improve the performance of medical staff has received great attention and is used in the health systems of different countries. The most important of these methods are Continuing Medical Education (CME), Community-Based Medical Education (CBME), and Continued Professional Development (CPD). We will investigate them in the following.

### 1- Continuing Medical Education (CME)

In the study of Wang et al. (2024), the needs of gynecologists in the field of education with CME methods and educational content were examined. Based on the findings, continuing education in the fields of female endocrinology, abnormal uterine bleeding, and polycystic ovary syndrome was needed by gynecologists and improve their performance in these fields (18). Jayas et al. (2023) noted that the need for physicians to stay up-to-date with medical information led to an increase in their willingness to participate in CME (19). Pott et al. (2021) noted that the topic, quality of content, availability of accredited CME, and focus on clinical practice were among the factors that influenced physicians’ choice of CME activities. Being live, online, and care-based were among the features that influenced physicians’ participation in CME. Because being online reduced costs and time, and being live led to better teaching and focused on content (20). Gill et al. (2016) evaluated the effectiveness of SMS-based CME on clinical knowledge retention of community-based physician assistants (CBPA). The findings did not show a significant increase in job satisfaction, self-efficacy, and medical knowledge scores (21). In a study by Shah et al. (2012), it was shown that the average weekly duration of self-study among Pakistani family physicians was 6.5 hours, and these physicians were trained without practice and acquired most of their knowledge from books. As a result, they emphasized the use of online CME to improve knowledge and performance (22). Liljestrand et al. (2004) showed that physicians and nurses providing health care to HIV patients who had participated in CME and had more experience in this field were more skilled and satisfied in providing counseling to patients and were more willing to recruit HIV patients (23). Davis et al. (1995) in their review study showed that the participation of physicians and medical residents in CME had a positive effect on health care. Effective change strategies included reminders, strategies to strengthen health care delivery including practical practice with patients, mediation interventions, information, and multimodal activities that could influence the training activities of physicians and medical residents(24, 25). Levinson et al. compared the effects of two short-term programs (a 4-hour workshop and a 2-hour course) and long-term programs (a 2.5-day course) on the communication skills of physicians with patients. Based on the findings, physicians who had participated in the long program asked more open-ended questions, asked more patient opinions, and provided more biomedical information. Also, their patients tended to provide more biomedical and psychosocial information, and by reducing negative affect between the patient and the doctor, patients tended to show fewer signs of external distress. Which ultimately increased their satisfaction (26). Eickmann et al. (2024) conducted a qualitative study in German hematologists that examined barriers and facilitators to participation in CME. Personal limitations, structural limitations, and content-related limitations were among the barriers to learning and training. To overcome these barriers, attention should be paid to identifying appropriate CME courses, targeting specific audiences, and training tailored to individual needs to improve performance (27). In general, it can be noted that the use of the CME method can improve the knowledge and performance of medical personnel, physician assistants, and nurses in the field of health services.

### 2- Continued Professional Development (CPD)

Continuing professional development (CPD) is a cornerstone of primary health care. Al-Sheikhly et al. (2023) emphasized the necessity of CPD to improve physician performance and mentioned that activities during working hours, cost, and work commitments are the most important barriers to physician participation in this program, which should be addressed to develop physician participation as much as possible. Participants also preferred workshops, traditional lectures, case-based sessions, small groups, and online learning to participate in CPD (28). Tavares et al. (2022) mentioned the use of experiential educational simulation for CPD as another method that can increase physician participation in CPD. Simulation is an experiential educational method that helps individuals experience a therapeutic method with a clinical intervention in a clinical and practical environment. As a result, using this method to train and improve physician performance can be an effective measure, although factors such as familiarity with simulation, psychological safety, and direct connection to clinical practice are effective in increasing participation (29). Kozikowski et al. (2021) noted that medical assistants seeking the National Council for Physician Assistant Certification (NCCPA) reported using CPD as an effective way to update their medical knowledge (30). In a study by Whicker et al. (2020), continuing education, career planning, legal requirements, and commitment to learning were among the factors influencing CPD. CPD was also shown to play an important role in the professional development of physicians (31). Breunig et al. (2020) showed that the use of clinical cases in CME for the training of physician assistants (PAs) and nurse practitioners (NPs) significantly improved the effectiveness of teaching in PA and NP learners (32). Alexander et al. (2018) showed that participation in a professional development program for PAs improved knowledge, skills, and resources. However, it was noted that educational success, program director stability, involvement in leadership, and service activities had a positive impact on participants’ professional development (33). Howie et al. (2017) stated that continuing professional development (CPD) is important for all healthcare professionals and improves performance and quality of patient care. As a result, it was recommended that educational supervisors use learning strategies to train PAs that enhance their clinical skills (34). Rao et al. (2010) in the UK emphasized the use of CPD to train physicians and psychiatrists to improve patient care, and good psychiatric practice and increase their credibility (35). CPD is one of the effective methods for training and developing the performance of medical personnel, as a result, studies have emphasized the use of this method to improve the knowledge and performance of this group.

### 3- HPE, CBME and RTME

RTME is another method that can have a positive impact on the knowledge, skills, and performance of physicians and the medical team. Kulo et al. (2022) reviewed that health professions education (HPE) can enhance the professional development of health professions faculty and can enhance their knowledge and skills for teaching and learning roles (36). Miskeen et al. (2023) reported that primary care physicians (PCPs) had a reasonable level of awareness and engagement in community-based medical education (CBME) classes and demonstrated that this perspective could facilitate curriculum development and create effective partnerships for CBME participation (5). Jin et al. (2023) found that rural tuition-free medical education (RTME) for free physician education led to increased physician job satisfaction, greater promotion opportunities, a relaxed work environment, the development of community health services, and easier access to higher technical titles (37). In general, using the aforementioned strategies, taking into account individual circumstances, and providing content and appropriate needs of physicians, can improve the knowledge and performance of physicians and medical assistants in providing health services.

## DISCUSSION

Education is the main pillar of acquiring knowledge and keeping it up to date. By acquiring knowledge, one can improve skills and improve performance. Given the continuous advancement of medical science and related knowledge, continuous acquisition of knowledge in this field is essential (38). Accordingly, various strategies, including CME, CPD, and CBME, are used to provide continuous education and educational content to medical students, physicians, and medical residents (39, 40). These strategies help these groups acquire knowledge by designing and making available the educational content needed by these groups so that many countries have made participation in these courses and educational content necessary and mandatory. The characteristics of these strategies include the appropriateness of the subject, method, and educational content of CME, keeping medical information up to date, improving the quality of content, accessibility of CME, focusing on clinical education, being web-based and online, increasing job satisfaction and self-efficacy, practical practice, and being care-based in services. The results of the studies reviewed in the present study have shown that the use of the aforementioned strategies can improve the knowledge and performance of physicians in providing health services (41, 42).

Physicians working in healthcare systems are under great pressure due to the rapid change and development of medical science, treatment methods, and healthcare(43, 44). Continuing medical education helps them to adapt to medical advances, familiarize themselves with them, and be able to provide optimal care (42). Medical education using CME, CPD, and CBME strategies and the frameworks determined by education officials in organizations helps physicians balance professional responsibilities and current knowledge and improve performance and efficiency in learning and providing quality services (45-47). In general, continuing medical education using the aforementioned strategies plays a fundamental role in the professional development of physicians. Medical education can significantly change their knowledge, attitudes, performance, and skills. The use of CME, CPD, and other valid, innovative, and flexible strategies has important effects in meeting clinical needs and healthcare services. These strategies can enhance the competence of physicians and provide valuable opportunities to improve the quality of services in hospitals and health centers (40, 48, 49).

In the present study, barriers such as inappropriate topics, content, time, time commitment, cost, and lack of financial support limited participation in continuing education programs. Studies have indicated that educators and professors involved in CME design should consider personalization, repeatability, and adaptability to advances in educational technology and develop educational programs to maximize participation (50). In general, using learner- centered activities and a wide selection of strategies and content areas to meet the needs of physicians in their living and working environments, and choosing learning methods such as webinars, conferences, group discussions, problem-solving, and case studies that can form part of the educational activities in the lives of physicians and be available to them. In general, most physicians need a combination of educational opportunities to access the best practices and to meet their educational needs by selecting appropriate educational activities. CME, CPD, and CBME strategies can be very helpful in this regard (51, 52).

## CONCLUSION

The present study showed that educational strategies including CME, CPD, and CBME can improve physicians’ knowledge and performance, increase job satisfaction, self-efficacy, communication skills, knowledge, performance, and medical skills, and create a positive change in the provision of health care. However, individual, structural, and content limitations were among the barriers to learning and training in this field, which need to be addressed to improve the use of these strategies and physicians’ performance.

## Data Availability

All data produced in the present work are contained in the manuscript

## Declaration

### Ethics approval and consent to participate

Not applicable

### Data Availability Statement

All data generated or analyzed during this study are included in this published article.

### Consent for publication

Not applicable

### Authors’ contributions

E.A. has conducted all parts of the study, including design, execution, and writing the manuscript.

## Acknowledgments

The author would like to thank the Research Vice-Chancellor of Shiraz University of Medical Sciences.

